# Health systems trust in the time of Covid-19 pandemic in Bangladesh: A qualitative exploration

**DOI:** 10.1101/2020.08.05.20157768

**Authors:** Taufique Joarder, Muhammad Nahian Bin Khaled, Shahaduz Zaman

**Affiliations:** Executive Director, Public Health Foundation, Bangladesh; Research Analyst, International Food Policy Research Institute (IFPRI), Bangladesh office; Reader in Medical Anthropology and Global Health (Global Health and Infection), Brighton and Sussex Medical School, University of Sussex, UK

**Keywords:** Covid-19, Pandemic, Trust, Resilience, Health policy and systems research, Bangladesh

## Abstract

**Background:** Lack of trust hinders care seeking, and limits community support for contact tracing, care seeking, information and communication uptake, multisectoral or multi-stakeholder engagement, and community participation. We aimed at exploring how trust might be breached and what implications this may have in Covid-19 pandemic response by the Bangladesh health systems.

**Methods:** We conducted this qualitative research during the pandemic, through seven online focus group discussions, with purposively selected mixed-gender groups of clinicians and non-clinicians (n=50). Data were analyzed through conventional content analysis method.

**Results:** The common thread throughout the findings was the pervasive mistrust of the people in Bangladeshi health systems in its management of Covid-19 pandemic. In addition to the existing health systems weaknesses, few others became evident throughout the progression of the pandemic, namely, the lack of coordination challenges during the preparatory phase as well as the advanced stages of the pandemic. This; compounded by the health systems and political leadership failures, lead to opportunistic corruption and lack of regulations; leading to low quality, discriminatory, or no service at all. These have trust implications, manifested in health seeking from unqualified providers, nonadherence to health advices, tension between the service seekers and providers, disapproval of the governance mechanism, misuse of already scarce resources, disinterest in community participation, and eventually loss of life and economy.

**Conclusions:** Health sector stewards should learn the lessons from other countries, ensure multisectoral engagement involving the community and political forces, and empower the public health experts to organize and consolidate a concerted health systems effort in gaining trust in the short run, and building a resilient and responsive health system in the long.

**Key Messages:**

1. **Implications for policy makers**
  - The preexisting health systems weaknesses, widely discussed in many literatures on Bangladeshi health systems, need to be addressed first, in consultation with health policy and systems experts.
  - In order to improve the coordination and science-based professional response to Covid-19 pandemic, the relevant experts, instead of administrators or bureaucrats, should be immediately engaged and deployed.
  - In order to facilitate adaptive leadership, health system should ensure transparency in every aspects of its functions, curb corruption and discrimination, regulate private sector for cost and quality of services, and ensure equity and fairness.
  - Politicians in power should engage with other social, cultural and religious forces and formally engage with other political parties in facing the Covid-19 crisis, with a view to fostering multisectoral collaboration and community engagement.
  - The health system actors should ensure a free flow of correct information following evidence based, scientifically oriented social and behavior change communication (SBCC) strategies.
2. **Implications for public** Bangladeshi health system is grappling with the Covid-19 pandemic. The authors believe that a better response was possible. In this research, people themselves expressed their perceptions and views regarding the alleged mishandling of the situation by health systems stewards. Careful addressing of the issues explored in this article may lead to a better pandemic response in the short run, and develop a resilient health system in the long.

## Background

In general understanding, ‘trust’ means, if trustors turn to trustees, the trustee would not exploit the trustor, rather, protect the vulnerable trustor from which s/he turned to the trustee^1-4^. For example, the Covid-19 pandemic has made people vulnerable to the disease, social relations, livelihood, etc. So, the health systems authority is expected to protect the people, and should not exploit them in the face of their vulnerability. Different levels of trust have been theorized depending on whether trust is in a person, a system or institution^2^. Sripad et al. (2018) described ‘trust’ in terms of interpersonal and impersonal trust in the context of maternity care in Kenya, where they defined the former as the trust between service provider and service seeker; and the latter as that in a social system, such as the health systems^5^. In this article, by ‘trust’ we refer to the impersonal trust, i.e., the trust of the people in the Bangladeshi health system during the Covid-19 pandemic.

Health systems are comprised of persons, institutions or activities whose primary intention is to improve people’s health^6^. Ministry of Health and Family Welfare (MoHFW) is responsible for policy, planning, and overall governance of the health sector through different directorate generals, which are responsible for policy implementation. Pandemic response is guided by Infectious Diseases (Prevention, Control and Elimination) Act 2018, which places Directorate General of Health Services (DGHS) as the central coordinating and responsible body for Covid-19 response. Under this; Institution of Epidemiology, Disease Control, and Research (IEDCR) is the main scientific body to provide technical guidance and support for screening at point of entry, quarantine, contact tracing, testing (initially, then later contracted out to other government and a few private laboratories), forecasting, surveillance, and outbreak response^7^. In the urban areas, however, Ministry of Local Government, Rural Development and Cooperatives is responsible for primary level healthcare, while secondary and tertiary care is bestowed upon an unregulated (or poorly regulated) private sector^8,9^. As of 7 July 2020, 168,645 cases (#17 in the world) have been identified with a death toll of 215^10^, 3rd highest new cases (3027) per day, and one of the lowest rate of tests per million (5,321) in the world and the second lowest among its South Asian neighbors (only above Afghanistan)^11^. Major events related to Covid-19 pandemic and its response in Bangladesh have been shown in Figure 1.

**Figure 1.**
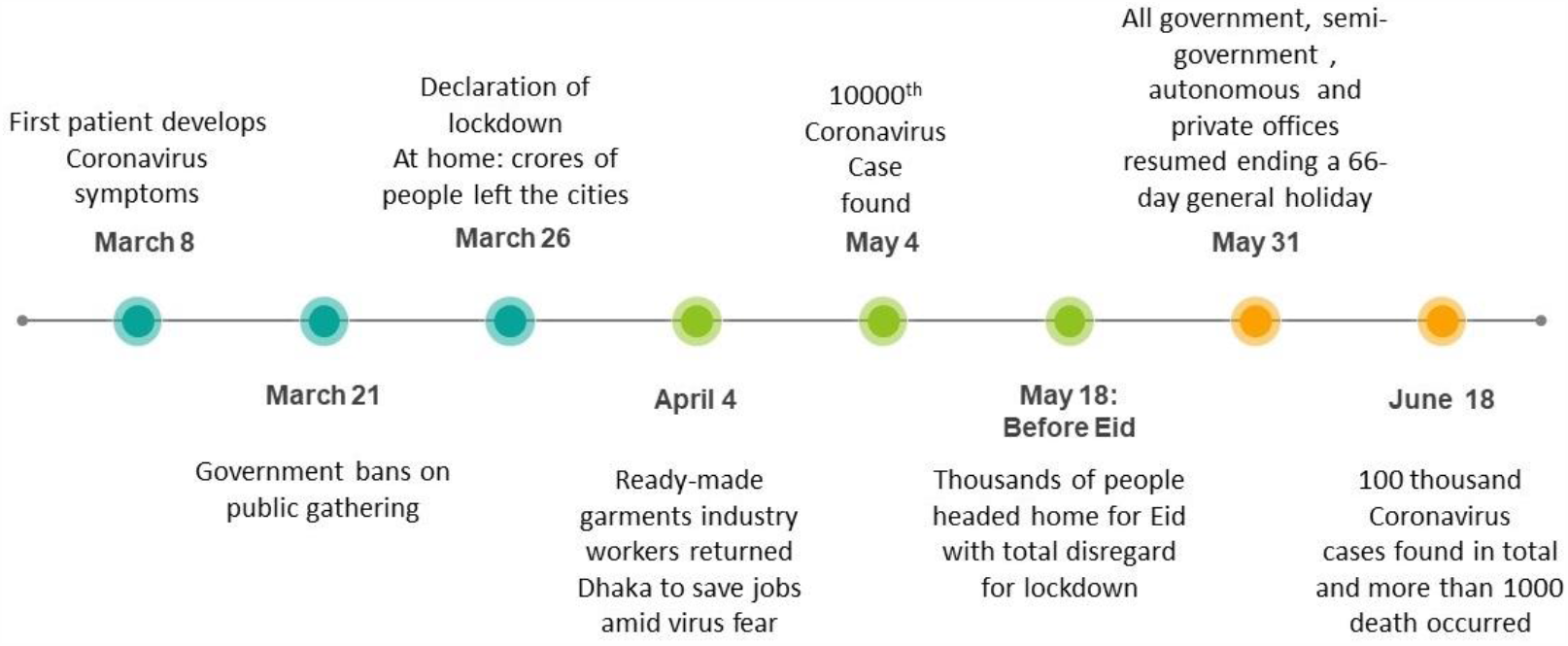
Covid-19 timeline in Bangladesh

Trust is important in health systems as it fosters cooperation among diverse stakeholders and contributes to health production^2^, which are imperative in responding to a pandemic in the short term and building a resilient health system in the longer term. It plays a critical role in garnering social order among the population^2,12^ and feeding back into the improvement of the governance as a whole^13,14^. It improves communication flows which are essential for risk communication to the people in a pandemic^15,16^, and information flows which are essential for technical and managerial decision-making by experts and managers^2,17^. Sharing information by cases and contacts (as part of contact tracing activities) is a cornerstone in pandemic response, which depends on public’s trust on the health systems and the state at large^2,15,17^. Lack of trust may invoke widespread stigma regarding infectious diseases, which severely obstructs case detection, isolation, quarantine, and case management activities^18,19^. People’s trust in health systems improves access, utilization, adherence, and continuity of care, leading to improved health and satisfaction^12,17,19^. Not only among the general people, but also among the service providers, trust is important, as it improves motivation and service provision; and lack of it can escalate to strike, e.g., against the lack of personal protective equipment (PPE) and medical equipment^20^ Finally, Covid-19 response requires adaptive leadership, i.e., making bold decisions and passing newer regulations with the emergence of newer evidences, which is impossible without trust between decisionmakers and the range of stakeholders and the people^21^.

However, while these examples suggest that trust is important in Covid-19 response and developing a resilient health system in the long run, understanding on trust in health systems context is ambiguous. There is only one systematic review on trust measures, and their content areas^12^, and a few other articles on trust and healthcare^2,3,5,22^. Most of the studies, however small in number, are from Western countries^14^. Role of trust in pandemic preparedness and response is rare globally^17^, and absent in Bangladesh. Therefore, this study aims at exploring how trust might have been breached and what implications this may have in regard to the Covid-19 pandemic response by the Bangladesh health systems.

## Methods

### Study design

This was an exploratory qualitative research, conducted during the lockdown situation of Covid-19 pandemic in Bangladesh, through online focus group discussions (FGDs), to obtain a deeper understanding of the construct of trust in the health systems context of Bangladesh.

### Study context

This research was sponsored by United Nations Youth and Students Association of Bangladesh (UNYSAB), which organized a webinar on 19 May 2020, amid the lockdown situation (officially termed as ‘General Holidays’) in Bangladesh, on ‘Trust in the Health System in COVID-19 Pandemic and Beyond.’ Moderated by the first author, the discussants included a public health expert from the not-for-profit private sector (Head of the communicable disease program of a leading non-governmental organization of Bangladesh), a public health expert from the public sector (an Assistant Director of a government health agency of Bangladesh), a health economist (an Associate Professor at a leading Bangladeshi public university), and a female entrepreneur. During the webinar, the moderator and the first author described the basic tenets of trust^12^ and asked the discussants about the role of trust in Covid-19 pandemic response, how trust may be breached, how breach of trust may impact the health systems, and what may be done to improve the trust in building a resilient health systems. This webinar helped in gaining a practical overview of the trust situation in Covid-19 crisis response by the Bangladeshi health systems. During the webinar a Google Forms link was circulated by the organizers to collect the name and contact information (name, email, phone number, Skype ID, educational background, and occupation) of interested FGD respondents.

### Data collection

We conducted seven FGDs through 15 to 17 June 2020, with the respondents who showed interest in the Google Forms circulated during the webinar. Each FGD, held and recorded through the video conferencing software Google Meet, consisted of 6-10 respondents, selected through purposive sampling method, from the Google Forms list. Each FGD was done with one of the following groups of mixed-gender respondents, categorized into two broad categories: 1) Clinicians: graduate students (all with medicine or dentistry as their undergraduate background) of public health of a private university, renowned public health experts (all with a medicine background), practicing clinicians (with either medicine or dentistry background); and 2) Non-clinicians: undergraduate students of different departments (management, marketing, botany, business, and pharmacy) of a public university, undergraduate students of public health of a public university, undergraduate students of food and nutrition department of a public university, service holders of different professions (executives, trainers, managers, and coordinators of public and private organizations).

The FGDs were moderated by the first author, who is a health policy and systems researcher with experience and expertise in qualitative research methods. The second author, trained in economics and experienced in qualitative research, assisted in note taking. The open ended FGD guide addressed three main questions: 1) How trust might have been breached during the Covid-19 pandemic, 2) What are the implications of breach of the trust, and 3) How trust may be restored and improved for a resilient health system. Each FGD, lasting from 60 to 105 minutes, was conducted in Bengali, the native language of the respondents and the researchers.

### Data analysis

The FGDs were immediately transcribed by the members of UNYSAB, the sponsor of this research. For analysis, we adopted conventional content analysis method^23^ which is appropriate in scarcity of existing literature on a phenomenon, as this method avoids using preconceived themes. We developed the inductive categories through the following steps: familiarizing with the data by listening to the records and reading the transcript, developing coding schema in MS Excel based on the three questions mentioned above, noting the first impressions followed by labelling the text segments by newly emerging codes, merging the similar-meaning codes together, then sorting the codes into larger categories based on how different codes are related or linked. Then we applied the concept mapping technique to organize these categories into a hierarchical structure (Figure 2). In order to increase validity, the first and second author independently coded the dataset, and the third author was involved where a third opinion was needed to reach a consensus.

**Figure 2.**
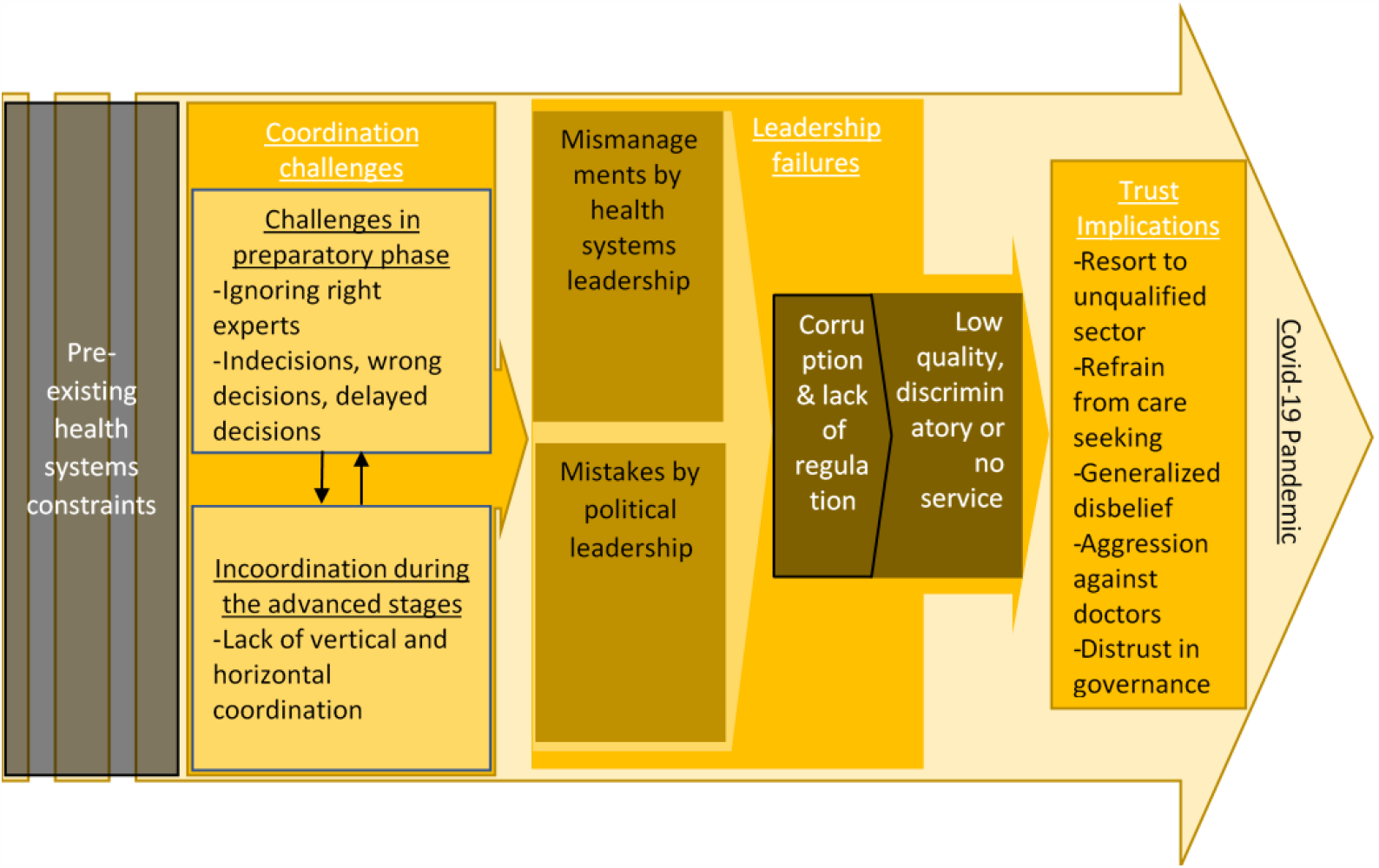
Model depicting health systems trust in Covid-19 pandemic in Bangladesh

## Results

### Background characteristics of the respondents

A total of 50 respondents participated in 7 FGDs (Table 1). Age of the respondents ranged from 19 years to 75 years, 28 were males and 22 females. 4 of the FGDs were held with respondents with non-clinical background (n=28) and 3 were with clinicians (n=22). Almost half of the respondents had training in public health.

**Table 1.** Characteristics of the FGD participants

| FGD Number | Group characteristics | Number of respondents | Age range in years | Male/ female | Clinical or non-clinical background |
| --- | --- | --- | --- | --- | --- |
| FGD-1 | Undergraduate students of different departments (management, marketing, botany, business, and pharmacy) of a public university | 6 | 19-21 | 5/ 1 | Non-clinical |
| FGD-2 | Graduate students (all with medicine or dentistry as their undergraduate background) of public health of a private university | 6 | 25-34 | 0/ 6 | Clinical |
| FGD-3 | Undergraduate students of public health of a public university | 9 | 21-26 | 4/ 5 | Non-clinical |
| FGD-4 | Undergraduate students of food and nutrition department of a public university | 6 | 22-25 | 0/ 6 | Non-clinical |
| FGD-5 | Service holders of different professions (executives, trainers, managers, and coordinators of public and private organizations) | 7 | 24-28 | 6/ 1 | Non-clinical |
| FGD-6 | Renowned public health experts (all with a medicine background) | 7 | 45-75 | 5/ 2 | Clinical |
| FGD-7 | Practicing clinicians (with either medicine or dentistry background) | 9 | 28-67 | 8/ 1 | Clinical |

### Breach of trust due to coordination challenges

Health system of Bangladesh has already been suffering from shortage of budget, lack of quality of services, and a highly centralized secondary or tertiary care. According to the respondents, Covid-19 just exposed the situation:

> *“The lack of trust in health systems predates the Covid-19 situation, and for a right reason. The pandemic simply exposed the fact that the emperor actually has no clothes*.*”* (FGD-6, renowned public health experts, clinical background)

Pre-existing inadequacies are compounded by the mistakes of the health system actors in the preparatory phase. Respondents observed several mismatches between what have been said and done. The health sector reportedly failed to place right persons in the right position in pandemic response, as expressed by a professor of public health:

> *“An epidemic is a public health emergency; it is neither clinical nor an administrative issue. So, we must see this problem through the public health lens. We* [epidemiologists] *already know what to do to control an epidemic. …We the public health professionals should be given flexibility that we are free to do whatever is needed for the country, not something that just pleases the political leadership*.*”* (FGD-6, renowned public health experts, clinical background)

Their lack of preparation is also evident from imposing home quarantine at the beginning of the pandemic, instead of an institutional one, while knowing that, intimate Bangladeshi culture is clearly not conducive to home quarantine. Insufficient testing, delay in giving test results, high cost of diagnosis and treatment in private sector, insufficient equipment in the health centers, etc. also indicate a lack of farsightedness among the health system actors:

> *“We are not getting the test results in time. My friend’s father got test report after 10 days, when he was already dead*.*”* (FGD-3, undergraduate students of public health of a public university, non-clinical background)

Respondents alto think that, not ensuring subsistence allowance for the poor before imposing a lockdown (official term was ‘general holiday’), not allowing people to take preparation as they did not mention how long the lockdown might continue, and increasing the duration week by week were signs of unreadiness, leading to ineffective lockdown. The health system actors wasted critical time before allowing private centers, although only a few in number, to provide Covid-19 care; conducting tests from only one government facility during the initial days of the pandemic, and not using alternative testing facilities like all the private and non-governmental organizations’ (NGOs’) reverse-transcriptase polymerase chain reaction (RT-PCR) and rapid testing facilities. A student of public health said:

> *“Government is allowing, although lately, private diagnostic centers to perform Covid-19 test. But there are many other RT-PCR machines which are not yet being utilized*.*”* (FGD-2, graduate students of public health of a private university, clinical background)

Lack of coordination among different government departments, between government and other sectors, and with different business agencies regarding whether or when those businesses will open, created a massive confusion. One such incoordination allegedly led to massive spread of the disease among the low-income garment employees:

> *“BGMEA* [national trade organization of garment manufacturers] *ordered garment workers to return to Dhaka to save their job. After they returned by thousands, braving the coronavirus infection, BGMEA declared that the factories will not open. They* (BGMEA) *never consulted public health experts or health department. On the other hand, the police announced that they will not allow anyone to enter Dhaka. The innocent workers were caught between a rock and a hard place, just because of the incoordination of different departments*.*”* (FGD-2, graduate students of public health of a private university, clinical background)

Lack of coordination was also observed in imposing lockdown in periphery areas of the country without arranging the transport of the patients to the centrally located health facilities, declaring several hospitals as dedicated Covid-19 hospitals while these were not prepared, asking private hospital to provide care while testing was not allowed for them to verify if they are admitting Covid-19 positive patients with general patients, suddenly opening markets before Eid vacation while health experts at the government committees were calling this an unscientific decision, etc. A respondent said:

> *“All the good things that we have in the health sector are based in Dhaka* [capital city]. *The health bureaucrats at district and sub-district levels must coordinate with the civil administration to ramp up the response against Covid-19. Success cannot be achieved without a tactful decentralization, involving other ministries as well*.*”* (FGD-6, renowned public health experts, clinical background).

### Breach of trust due to leadership failures

The health systems made several good decisions, but those also faced criticisms for poor management. For example, point of entry screening was weak from the beginning, government officers were seen putting on personal protective equipment (PPE) meant for the doctors and posing on television, etc. Not receiving the PPEs in time made many doctors diagnosed with Covid-19, and this created a panic among the general people:

> *“If doctors are getting affected so much, and they are not getting care, then how can we get care?”* (FGD-4, undergraduate students of food and nutrition department of a public university, non-clinical background)

Miscommunication was another action that attracted much criticism. For example, conflicting statements from different responsible persons, opening businesses only to change the decision multiple times without clear directions, and declaring lockdown in the guise of ‘general holiday’ that was extended week by week:

> *“When they (government) say it is a ‘general holiday’ instead of ‘lockdown’, people confuse it with something festive. That’s why we saw people going to Cox’s Bazar* [a popular tourist destination] *for tourism purpose. Even some of my friends got married, taking advantage of the ‘general holiday’*.*”* (FGD-3, undergraduate students of public health of a public university, non-clinical background)

Apart from the health systems actors, the political leadership of the country are believed to downplay the risk and added fuel to the fire by irresponsible remarks. A popular perception is that the government procrastinated in ramping up the Covid-19 response in the early days of the pandemic with an intention to make some political programs hassle-free. There is an impression that the government put political agenda above health agenda, which allegedly became evident by the incident of disapproving an antibody-based testing kit developed by a local NGO, due to the alleged political affiliation of the leader of that NGO with the opposition party. A doctor said:

> *“We are saying antibody testing is essential, they* [the government] *are saying no. Are they doing it politically!* (FGD-7, practicing clinicians, clinical background)

It is also believed that the private testing facilities and hospitals that are getting approval lately have political blessings. A student of public health said:

> *“There is a rumor that government is allowing diagnostic centers those are politically close to them. For example, Popular Diagnostic Center* [a private diagnostic company, rumored to have ties with an opposition political party] *is denied to conduct tests due to political reasons. This is not the time for politicization*.*”* (FGD-2, graduate students of public health of a private university, clinical background)

Other criticisms against the political response include positioning the rhetoric of livelihood against life under the pressure of business leaders, giving false hope repeatedly while knowing that they were not prepared, and defending themselves against their failures without confessing their mistakes or apologizing, etc. A respondent said:

> *“If a leader confesses his mistakes, I may be angry at first, but my trust will return, because, he is telling the truth, at least*.*”* (FGD-5, service holders of different professions, non-clinical background)

Incidents of several corruptions made to the media, many of which are perceived to be linked with politics. For example, many PPEs were found to be counterfeit yet got approved by the health department, several decisions to protect the interest of the businessmen were seen as an offshoot of political consociation, etc. Lack of regulation was another complaint, examples of which include: high price of essentials due to panic buying, mushrooming of unauthorized and even fake testing centers, uncontrolled advertisement and selling of putative Covid-19 medicine (e.g., hydroxychloroquine, ivermectin, remdesivir, etc.), and lack of action against the false Covid-19-negative certifications:

> *“Today I saw in the news that, someone who had been found corona negative here, was found positive after landing in Japan. So, when things like these happen, it calls upon our trust. These instances may even adversely affect our foreign relations*.*”* (FGD-5, service holders of different professions, non-clinical background)

Trust has been breached by low quality services due to scarcity of essential facilities like intensive care unit (ICU) beds, equipment like ventilators, oxygen supplies, etc. These are supplemented by the information that private hospitals are denying to admit patients without Covid-19-negative certificate; obtaining which takes long time due to lack of adequate testing facilities, eventually deteriorating the condition of the patients. Those with respiratory symptoms suffered the most, leaving several patients dead in the process of back and forth between multiple hospitals. These allegations were compounded by the rumors of private hospitals keeping patients admitted for more money even after getting cured, charging astronomical amount of money for scarce services like ICU or oxygen, taking patients hostage for money, etc.:

> *“My relative was kept in a private hospital for extra three days after she was found covid negative. She was even given oxygen. When asked why, they said it is for extra safety of the patient*.*”* (FGD-3, undergraduate students of public health of a public university, non-clinical background)

Discriminatory care was the worst allegations against the health system. There are circulating stories of high profile people booking the whole hospital for their family members, leaving country of some business tycoons by air ambulances or chartered flights, and the alleged reservation of a public hospital only for the so called ‘very-important-persons’, known popularly as VIPs:

> *“We are seeing that wealthy and political people of the ruling party are getting one type of treatment, while general people are getting something different. Some managed to go out of the country by chartered flight, some ministers booked ICUs even before requiring one. Hearing such news, as a middle- or lower-middle-class member of the society, I can’t help losing trust in the health system. All facilities are there to protect the upper layer of the society*.*”* (FGD-4, undergraduate students of food and nutrition department of a public university, non-clinical background)

Taking service by government health sector high officials not from public sector hospitals but from a well-known military hospital instigated much mistrust among the people, as expressed by a physician:

> *“When the top government health officials take treatment from the Combined Military Hospital, how can others keep trust on the government health system? What can be more painful than this?”* (FGD-7, practicing clinicians, clinical background)

### Implications of breach of trust in the health system

As a result of longstanding pre-existing health systems constraints, people will resort to popular or folk sector, especially with rising popularity of app- or web-based self-medication options. Seeking treatment from abroad is already a popular tendency, which will increase, leading to further loss of foreign currency:

> *“High income group will go abroad for simple cases, low income group will go to unqualified providers, middle class people like me will have nowhere to go*.*”* (FGD-2, graduate students of public health of a private university, clinical background)

As a result of lack of proper preparation and coordination, people would refrain from seeking care, i.e., they will not get tested, go to health facilities, maintain quarantine, self-isolate if suspected of or diagnosed with Covid-19, and eventually further spread the disease:

> *“General people like us will develop hesitation in their mind to seek healthcare. They will hesitate to get tested, which is the first step after suspecting Covid-19. They will not know whether they are infected or not and eventually infect their family members*.*”* (FGD-1, undergraduate students of different departments of a public university, non-clinical background)

Ineffective lockdown and other missteps sparked a negative perception and suspicion among the general people which made them disbelieve even the correct information and government directives. People’s disbelieves range from the number of Covid-19 positive cases reported by the government to the information shared on social distancing and other hygiene measures. A respondent said:

> *“When the schools were closed, my mother went to Kutubdia* [a distant island, respondent’s ancestral home] *with my siblings. I heard from her that people of Kutubdia are not even believing that any such disease even exists in reality. They think it’s a hoax, or a propaganda being spread by the government for a clandestine purpose. People are not abiding by any health messages delivered by the government*. (FGD-5, service holders of different professions, non-clinical background)

Loosing trust on the doctors, people may not adhere to the doctor’s advice, health messages, and treatments. People consider doctors or service providers as the representative of the health system, although, according to the FGDs with the clinicians, they are none but the victims of the system itself. But people often show aggression against the doctor; even a doctor was recently murdered by a dead patient’s aggrieved relative:

> *“A doctor has been beaten to death, but nothing has happened yet. …We the doctors always bear the burden of breach of health systems trust*.*”* (FGD-7, practicing clinicians, clinical background)

People will lose trust on the overall governance and the political system of the state, as a result, even the positive achievements will be disapproved by the people, demotivating the political leadership:

> *If I go to a shop and ask for good quality product, if I don’t have trust in him, then even if the shopkeeper gives a good product, I will not be satisfied with it. Government had achieved some praise by reducing the maternal mortality, increasing the life expectancy. Now that many people are dying of Covid-19, I will not trust the government despite their past successes*.*”* (FGD-5, service holders of different professions, non-clinical background)

Examples of corruptions, lack of regulations, and discriminations will push people further towards corruption, panic buying, overwhelming the testing facilities, health centers, etc. One respondent said:

> *“Since people don’t have trust, those who have scopes, just get tested even when it is not indicated, as if they are doing it as a hobby, at the cost of those who are genuinely in need of it. I will not be surprised to find some people who get tested even everyday, just because they afford it. Similar thing is happening with the hospital beds. We know, not everyone needs to get admitted, such as those who are having mild symptoms. I know people who, despite having mild symptoms, are occupying hospital beds unnecessarily. Why? Because they are afraid that they may not find a hospital bed a few days later if such a need arises. Even some people, who are not sick, have just booked cabins in case they need it sometime in the future. These could be avoided, had we been able to convince people that they will get necessary healthcare in time of emergency*.*”* (FGD-6, renowned public health experts, clinical background)

Negative perceptions and suspicion towards the system is a perfect recipe for stigma, mental health conditions, lack of community interest and engagement for a collective response. A respondent said:

> *“People feel insecure which may have a negative impact on mental health. People are so insecure that it has made them inhuman too. We heard that a son has left his mother in a jungle when she developed symptoms of Covid-19. We ourselves are experiencing the effects of stigma too, as many physicians are driven out of their homes by their landlords fearing spread of infection to their apartment*.*”* (FGD-2, graduate students of public health of a private university, clinical background)

Psychological impact of the whole failure would frustrate the people, and young people will leave the country. Both health and economy of the country will fall inside a positive feedback loop. A professor of public health said:

> *“We often say that public health is about saving lives in millions. But a blunder in public health can jeopardize the lives of millions too*.*”* (FGD-6, renowned public health experts, clinical background)

A summary of the findings and the recommendations are given in Table 2.

**Table 2.** Summary of findings and recommendations

| How trust might have been breached | Implications of breach of trust | Recommendations |
| --- | --- | --- |
| **Pre-existing constraints** |  |  |
| Insufficient budgetary allocation in health<br>Lack of quality of care<br>Highly centralized secondary and tertiary care | Self-medication<br>Resorting to unqualified service provider<br>Seeking medical care from abroad<br>Loss of foreign currency | Increase budgetary allocation and efficiency<br>Improve quality of care by enforcing quality standards and guidelines<br>Hiring varied workforce for improved skill-mix<br>Decentralize testing and healthcare |
| **Coordination challenges** |  |  |
| **Preparatory phase**<br>Right persons not in right place<br>Imposing home quarantine, instead of institutional<br>Insufficient testing<br>Delay in getting test results<br>High cost of care and testing in private sector<br>Insufficient equipment at health centers<br>Ignoring subsistence of poor before lockdown<br>Not allowing people to get prepared for lockdown<br>Not specifying duration of lockdown and increasing time week by week<br>Delayed approval of private sector to provide care<br>Not utilizing all testing facilities in the country from the beginning<br>Delayed decision in using rapid testing<br>**Incoordination during the advanced stages**<br>Incoordination among government departments | People refrain from seeking care<br>Will not get tested<br>Will not maintain quarantine<br>Will not self-isolate if diagnosed<br>Further spread the disease as a consequence<br>Negative perception and suspicion among general people<br>Ineffective lockdown<br>Disbelieve even in correct information by the government<br>Ignore government regulations and directives<br>Ignore the hygiene messages<br>Nonadherence to doctor's advices, health messages and treatments<br>Aggression against doctors | Introduce separate public health track<br>Scientific instead of administrative response<br>Ensure multisectoral collaboration<br>Introduce rapid testing kit along with RT-PCR<br>Improve environment of testing facilities<br>Impose lockdown after subsistence support<br>Communicate information on duration and alternative arrangements before lockdown<br>Coordination with private sector<br>Improved coordination among ministries<br>Ensure services and inform what |
| Incoordination between government and other sectors<br>Incoordination with business sector regarding opening businesses<br>No arrangement for emergency patients transport in lockdown<br>Dedicating unprepared hospitals for Covid-19<br>Asking private hospital to give care without allowing diagnostic tests<br>Sudden opening of markets before Eid |  | services are available and where |
| **Leadership failures** |  |  |
| **Actions by health system leadership**<br>Weak point-of-entry screening<br>Delayed distribution of PPE to doctors<br>Conflicting statements from high officials<br>Lack of clear guideline for opening businesses<br>Declaring ‘general holiday’ in the guise of lockdown<br>Increasing lockdown week by week<br>**Actions by political leadership**<br>Downplayed the risk<br>Irresponsible remarks by politicians<br>Prioritized political celebration over health<br>Political agenda before health agenda<br>Private services granted based on political affiliation<br>Rhetoric of life versus livelihood under the pressure of business leaders<br>Giving false hope, while knowing the leadership is actually unprepared<br>Defending mistakes, not confessing<br>**Corruption and lack of regulation** | Existing doctors will be demotivated<br>People will lose trust on overall governance mechanism of the country<br>Disapprove positive achievements of the past<br>Further corruption<br>Panic buying<br>Overwhelming testing facilities<br>Advance booking of health facilities<br>Stigma<br>Mental health conditions<br>Lack of community participation<br>Frustrated youth will leave country<br>Vicious cycle of ill health and poor economy | Improve multisectoral collaboration and coordination<br>Restrict misinformation and miscommunication<br>Ensure evidence-based management guidelines<br>Use clear, culturally sensitive, context specific terms supported by scientifically oriented SBCC strategy<br>Put public health before political priority<br>Regulate private sector for cost and quality<br>Confess mistakes after coming out of denial<br>Dissolve unholy syndicates in health sector<br>Take punitive actions against wrongdoers |
| Counterfeit PPE approved<br>Lack of regulation against panic buying<br>Mushrooming of fake testing centers<br>Advertisement of unproven drugs<br>False Covid-19-negative certification<br>**Low quality, discriminatory, or no service**<br>Scarcity of ICU facilities and equipment<br>Private hospitals denying patient admission<br>Delay in getting Covid-19-negative certificate<br>Death of many patients searching hospitals<br>Misdeed allegations against private hospitals<br>Discriminatory service for rich and powerful<br>Booking whole hospital by rich people<br>Business tycoons fleeing by chartered flights<br>Alleged reservation of public hospital for VIPs<br>Government health officials admitting in private hospitals for treatment |  | Free flow of evidence-based information<br>Effective communication channel up to rural<br>Messaging by religious and community forces<br>Policy and guideline against social stigma<br>Financial and nonfinancial motivation for service providers<br>Resolve professional grievances of service providers<br>Monitor and supervise health and other relevant staff<br>Ensure patients are not discriminated<br>Form all-party national consensus |

## Discussion

The common theme throughout the qualitative findings was the pervasive mistrust of the people in Bangladeshi health systems in its management of Covid-19 pandemic. This mistrust arguably stemmed from the existing health systems weaknesses, supplemented, throughout the progression of the pandemic, by, the lack of coordination challenges during the preparatory phase as well as the advanced stages of the pandemic. This; compounded by the health systems and political leadership failures, lead to opportunistic corruption and lack of regulations; leading to low quality, discriminatory, or no service at all. These have trust implications, manifested in health seeking from unqualified providers, nonadherence to health advices, tension between the service seekers and providers, disapproval of the governance mechanism, misuse of already scarce resources, disinterest in community participation, and eventually loss of life and economy. Understanding of these findings is important in reorganizing, restructuring, and revamping Bangladesh’s response towards Covid-19 pandemic.

One of the predictors of mistrust in Covid-19 response was the lack of coordinated actions. Many respondents, especially those with a public health background indicated that, this happened, because appropriate experts were not engaged since the early days of the pandemic. This is supported by the news articles and reports from Bangladesh. Bangladesh detected the first Covid-19 case on 8 March and experienced the first death on 18 March, but it formed the 17-member National Technical Advisory Committee (NTAC) On 19 April 2020. Until then, most of the pandemic control efforts were taken by the administrators, many of whom lacked expertise or experience in health, let alone pandemic management. Even the NTAC was devoid of adequate public health representation; among the 17-member committee, only three had a public health career track, others were high-profile clinicians such as neonatologist, endocrinologist, obstetrician, gastroenterologist, otolaryngologist, internal medicine specialist, anesthesiologist, psychiatrist, and so on^24^. On 21 April, the government made another announcement on assigning 64 top bureaucrats to supervise and coordinate relief distribution activities in 64 districts of Bangladesh^25^. The professionals were ignored once again in technical leadership for pandemic control, both at the central government and the local (i.e., district or sub-district) levels. A policy analysis on the human resources for health in Bangladesh also revealed that the DGHS is principally managed by the clinicians at the expense of public health experts. At the MoHFW level, the top health-bureaucrats are mostly drawn from other ministries, often unrelated to the health sector^26^.

Another theme that emerged as the determinant of health system trust was leadership, both in the health system and the political sphere. Ramalingam et al. (2020, p.2) argued, addressing a poorly understood and rapidly evolving disease like Covid-19 demands ‘adaptive leadership’, which cannot be achieved by “a small number of people behind closed doors.”^21^ They further added, implementation new policies and changing them as needed require candid and trustful relationships between policymakers and different stakeholders. This, however, is not the usual modus operandi in many countries, and our data indicates, Bangladesh is one of those countries. Participation, transparency, fairness, and trust-building—these ‘soft capacities’ are now considered as inseparable part of modern health system governance^27^.

Corruption, lack of regulation, rent seeking behavior by the health systems and political actors came out as dominant feature during the pandemic control. Bangladeshi newspapers support this perception of the people, by publishing articles on corruptions by companies or persons with alleged ties with the ruling political party. Such corruptions include, but are not limited to, granting permission for Covid-19 test to lesser known institutions which delivered false reports, approving low quality health products from corrupt companies, etc ^28-30^. A study by Transparency International identifies major health sector irregularities, which include politically influenced recruitment, transfers and promotions of healthcare professionals, irregularities in the procurement of drugs and equipment, unregistered and unqualified doctors operating in private healthcare facilities, and absenteeism^31^. A recent article, drawing examples from the Ebola crisis in Africa, demonstrated, how corruption erodes trust in the health system to the detriment of overall population health^32^.

It was evident from the remarks of the respondents that the lack of coordination and leadership in pandemic response led to mistrust and subsequent consequences, such as social stigma, psychological impact, and further spread of the disease. Zaman et al. (2020) in their rapid anthropological study, found that the information provided by the health system carried a varied meaning for the general people, both in urban and rural Bangladesh. The terms widely circulated by the media and the health authorities, such as quarantine, social distancing, lockdown, etc. were seen as foreign, ambiguous, and difficult to fit to the general Bangladeshi people’s context. The health messages thus circulated by several government and non-government organizations, largely in an uncoordinated manner, failed to synchronize with people’s social and economic reality, level of understanding, and risk perceptions^33^. Apart from noncompliance, different recent studies reported mistrust in the health system leading to widespread social stigma, fear, discrimination, rumor^34^, as well as psychiatric conditions^35^, even suicide^36^.

### Policy recommendations

The preexisting health systems issues are widely discussed in many literatures on Bangladeshi health systems, especially in ‘Bangladesh Health Sector Profile’ by the World Bank; and ‘Bangladesh Health System Review’^37^ and ‘Joint External Evaluation of IHR Core Capacities’ by the World Health Organization (WHO)^38^. These well-known health system weaknesses need to be addressed first, in consultation with health policy and systems experts. Next, to improve the coordination and science-based professional response to Covid-19 pandemic, the relevant experts for pandemic management should be immediately engaged and deployed. Such experts may include; but not limited to; public health professionals; including infectious disease epidemiologists, health policy and systems experts, medical anthropologists, health economists, health communication experts; laboratory scientists, including virologists, microbiologists, biochemists, lab technicians; relevant clinicians; including physicians, nurses, paramedics; and not merely administrators or bureaucrats. In the long run, however, a separate public health track in the health sector is indispensable^26^.

In order to facilitate an adaptive leadership, health system should ensure transparency in every aspects of its functions. To curb corruption and discrimination, it is important to take punitive actions, dissolve the unholy syndicates, ensure accountability, regulate private sector for cost and quality of services, and make sure no service provider can ever deny giving healthcare to any patient irrespective their social, economic, political, religious, gender, or any other kind of identity. Politicians in power should engage with other social, cultural and religious forces and formally engage with other political parties in facing the Covid-19 crisis, with a view to fostering multisectoral collaboration and community engagement^20,27,39^.

Finally, we recommend to ensure a free flow of correct information following evidence based, scientifically oriented social and behavior change communication (SBCC) strategies; restrict spreading misinformation; establish an effective communication channel up to the rural level; engage religious, cultural, political, and community based forces in message delivery; and formulate and implement an explicit policy and guideline to promote a culture of respect to counteract social stigma against Covid-19. WHO published the risk communication and community engagement (RCCE) action plan, where they discussed about regular and proactive communication with the public and at-risk populations to reduce stigma and build trust for the people and their families^40^. Information should be clearly and widely circulated for different groups, such as, the villagers, slum-dwellers, township residents, urban middle-class, etc. through contextually appropriate communication channels.

### Research implications

Since this study explored only the impersonal trust perception of the people in health system, a separate study my explore interpersonal trust, i.e., the trust between the people and the service providers. Secondly, a study on health systems governance may be conducted to obtain both the perspectives of the government actors as well as that of the other stakeholders outside the government. Patients’ perspective of the responsiveness of the service providers, and service providers’ perspective on their own safety and experiences of Covid-19 response could be other interesting areas to explore.

### Limitations

There are limitations to this study, such as, we could not achieve methodological or source triangulation due to the constraints of the pandemic situation. Secondly, the first author himself was a Covid-19 patient during the whole period of conducting this research. Given the sufferings of the disease and frustration of increasing rate of infection in the country, it is not unlikely to interpret the qualitative findings in a negative light. However, we tried to compensate for this by triangulation through multiple analyses^41^, i.e., by engaging multiple research team members in coding the data and interpreting the findings. Another limitation is that, since we had to conduct the FGDs online, we could not capture the perceptions of those who do not have access to internet, i.e., the less disadvantaged group, who may have less trust in the health systems. Also, we failed to capture the perspectives of the health systems policy or governance actors who are dealing with the Covid-19 pandemic on the ground. Therefore, it is important to keep in mind that this study reflects only the perceptions of a specific section of the society, and may not reflect the actual situation of health system governance in responding to Covid-19 pandemic.

## Conclusions

Pandemics have been known to the humans since the beginning of the recorded history, and perhaps even predate^42,43^. Covid-19 may be the first documented coronavirus disease^44^, but may not be the last. Bangladesh experienced several local disease outbreaks over the past several years^45-48^ and a recent dengue epidemic in 2019^49^. However, due to their lower magnitude compared to the Covid-19 pandemic, the need for a comprehensive overhauling of the health systems had not been felt so deeply. Low- and Middle-Income Countries like Bangladesh are particularly vulnerable to pandemics due to their week governance and health systems preparedness^27^. Therefore, a resilient health system is indispensable. The concept of ‘resilient health systems’ gained traction during the 2014-2015 Ebola epidemic in West Africa. Experts considered resilient health system as the precondition for combating future epidemics effectively. Resilience means the degree of change a health system can endure while maintaining its functionality^17^. A common thread in the resilience literature is the centrality of ‘trust’ in building resilient health systems^15^. In this article, we explored the pathways of breach of trust in health system during a pandemic situation. We realized many of the mistrusts stemmed from the already fragile and unprepared health system of Bangladesh compounded by lack of coordination and adaptive leadership. Based on our findings we recommend our health systems leaders to introspect and learn from the mistakes, in order to prevent a longer-term loss of life and economic downturn. We also feel that, our health sector stewards should take advantage of the lessons from other countries, ensure multisectoral engagement involving the community and political forces, and empower the public health experts to organize and consolidate a concerted health systems effort in gaining trust in the short run, and building a resilient and responsive health system in the long.

## Data Availability

We conducted 7 FGDs through 15 to 17 June 2020, with the respondents who showed interest in the Google Forms circulated during the webinar. Each FGD, held and recorded through the video conferencing software Google Meet, consisted of 6-10 respondents, selected through purposive sampling method, from the Google Forms list. Each FGD was done with one of the following groups of mixed-gender respondents, categorized into two broad categories: 1) Clinicians: graduate students (all with medicine or dentistry as their undergraduate background) of public health of a private university, renowned public health experts (all with a medicine background), practicing clinicians (with either medicine or dentistry background); and 2) Non-clinicians: undergraduate students of different departments (management, marketing, botany, business, and pharmacy) of a public university, undergraduate students of public health of a public university, undergraduate students of food and nutrition department of a public university, service holders of different professions (executives, trainers, managers, and coordinators of public and private organizations).

